# MRI Biomarkers for Brain Metastases in Lung Cancer Using Machine Learning

**DOI:** 10.64898/2026.10.12.26365323

**Authors:** Saahil Chadha, Durga V. Sritharan, Rohan Sharma, Darin Dolezal, Sampada Chande, Gregory Breuer, Daniel Fu, Arihan Gupta, Jahid Omar Hossain, Anand Srinivasan, David Weiss, Veronica L. Chiang, Don X. Nguyen, Sanjay Aneja

## Abstract

**Purpose:** Existing prognostic models for brain metastases from lung cancer rely on clinical variables and do not capture the biological heterogeneity of host or tumor factors underlying survival outcomes. We evaluated whether MRI-derived radiomic biomarkers improve prognostic stratification beyond clinical variables. We also investigated their biological correlates using matched histopathology.

**Experimental Design:** This retrospective study included 103 patients with brain metastases from lung cancer and matched pre-operative MRI and histopathology. Radiomic features from T1-weighted contrast-enhanced and T2-weighted fluid-attenuated inversion recovery sequences were combined with clinical variables to train Cox proportional hazards, random survival forest, and DeepSurv models for overall survival (OS), CNS progression, and leptomeningeal (LM) progression. Risk groups from an imaging-only model were compared for clinical and histopathologic differences.

**Results:** Imaging features improved prognostic performance beyond clinical variables across all modeling frameworks, with the combined clinical-imaging DeepSurv model achieving the highest concordance index for OS (0.728). Risk groups from the best-performing model for each endpoint differed significantly in OS (median 26.7 vs. 9.1 months, *p* < 0.001) and LM progression (*p* = 0.003), but not in CNS progression. Imaging-only risk groups did not differ significantly in any clinical variable, but high-risk patients were less likely to have tumor-infiltrating lymphocytes (46% vs. 77%, *p* = 0.010) and PD-L1 expression ≥ 50% (10% vs. 32%, *p* = 0.049).

**Conclusions:** MRI radiomic biomarkers provide prognostic information beyond established clinical variables in lung cancer brain metastases and correspond to features of the tumor immune microenvironment, supporting their potential to refine risk stratification.

**Statement of Translational Relevance:** Brain metastases affect 30-40% of patients with lung cancer. Existing prognostic models rely on clinical variables, which do not fully capture the biological heterogeneity that drives outcomes. We show that machine learning models integrating MRI-derived radiomic features with clinical variables improve prognostic discrimination beyond clinical variables alone. Because these biomarkers are derived from standard-of-care pre-operative imaging, they can be generated without additional testing or delay to treatment planning. Using matched histopathology from resected lesions, we further show that imaging-defined risk groups do not simply reproduce established clinical risk factors but instead correspond to differences in the tumor immune microenvironment, including tumor-infiltrating lymphocytes and PD-L1 expression. These findings support the use of quantitative imaging biomarkers as a non-invasive method to refine risk stratification at diagnosis, informing treatment selection and clinical trial enrollment for patients with brain metastases from lung cancer.

## Introduction

Brain metastases represent a growing clinical challenge, affecting 30-40% of patients with lung cancer and driving significant morbidity and mortality (1,2). Advances in the treatment of brain metastases have expanded the therapeutic landscape to include surgical resection, stereotactic radiosurgery, and advanced systemic agents including targeted therapies and immune checkpoint inhibitors. These treatment advancements along with the rising incidence of brain metastases have made management increasingly complex (3–5). There is an urgent need for prognostic models to stratify risk and guide treatment decisions that balance efficacy against toxicity.

The most widely accepted prognostic models for brain metastases have relied solely on clinical variables, like performance status and number of metastases, to stratify outcomes. More recently, it has been recognized that incorporating genomic data, such as driver mutation status, can improve prognostication (6–8). Although current prognostic models have been valuable for clinical decision-making, outcomes within risk strata remain heterogeneous, limiting their prognostic utility. This heterogeneity likely reflects underlying variation in host or tumor biology, including differences in microenvironmental composition, tumor morphology, and associated tumor invasion patterns that are not fully captured by clinical variables and mutation status alone (9,10).

Routine diagnostic imaging represents a rich and largely untapped source of biologically informative features that may address the heterogeneity within existing prognostic risk strata (11,12). While MRI is fundamental to the detection, surveillance, and therapeutic planning of brain metastases (13,14), imaging data has historically been excluded from prognostic models due to concerns over the subjectivity of qualitative image interpretation. Advances in quantitative imaging methods, particularly radiomic analysis, have mitigated these limitations by facilitating the extraction of quantitative features, including tumor morphology, intensity, and textural heterogeneity (15,16). By utilizing machine learning (ML) to integrate these high-dimensional data from standard T1-weighted contrast-enhanced (T1CE) and T2-weighted fluid-attenuated inversion recovery (FLAIR) MRI sequences, it is possible to model complex, nonlinear associations with clinical outcomes (17). Although such frameworks have improved risk stratification in various malignancies outside the brain (18,19), their application to lung cancer-associated brain metastases remains under-explored. Few imaging-based prognostic models have reached clinical use. Most have not been benchmarked against established clinical risk scores, so their added prognostic value over current practices remains unclear. In addition, these models are typically developed as statistical predictors, and relatively few studies have examined whether the resulting risk scores are associated with underlying tumor biology. Without biological correlation, it is unclear what the clinical risk scores capture, which limits trust and adoption. Lastly, current models focus almost exclusively on overall survival (OS) and do not assess prognostic value for other clinically relevant endpoints, such as CNS progression and leptomeningeal (LM) disease, which are arguably more clinically actionable.

To address these gaps, we evaluated the prognostic value of ML-derived MRI biomarkers in patients with lung cancer-associated brain metastases and benchmarked these biomarkers directly against a widely used clinical nomogram. Using three complementary survival modeling frameworks, we assessed the prognostic utility of clinical, imaging, and combined clinical-imaging models. Lastly, we assessed whether our ML-derived MRI biomarkers correlated with tumor immune microenvironment features to understand their biological basis for improved prognostication.

## Materials and Methods

### Dataset

We used data from an institutional biorepository of central nervous system metastases (Figure 1A). This study was approved by the Yale University Institutional Review Board (protocol 2000027592) and conducted in accordance with the Declaration of Helsinki, with a waiver of informed consent. The cohort included 111 resected brain metastases from 103 patients with primary lung cancer, each with a matched pre-operative MRI and histopathologic specimen, including 8 patients who underwent repeat resection and contributed both specimens. Pre-operative T1CE and/or FLAIR MRI sequences were obtained. Demographic and clinical variables obtained from the institutional dataset included age, sex, Karnofsky Performance Status (KPS), number of brain metastases, presence of extracranial metastases, histologic subtype, and epidermal growth factor receptor (EGFR) mutation and anaplastic lymphoma kinase (ALK) rearrangement status (Table 1; Figure 1B).

**Table 1.** Baseline demographic, clinical, and histopathologic characteristics of the study cohort, stratified by histologic subtype. The cohort consisted of 103 patients contributing 111 resected BM with matched pre-operative MRI. Demographic and clinical variables (age, sex, KPS, number of BM, ECM, EGFR, and ALK status) are reported per patient (n = 103). Lesion size, PD-L1, and TIL density are reported per resected lesion (n = 111). Percentages are within each histologic subtype. PD-L1 TPS was obtained from clinical pathology reports. TIL density was graded semi-quantitatively by a board-certified pathologist on H&E-stained sections of the resected lesion, and was dichotomized as absent (0) versus present (1+ or 2+) for analysis. Continuous variables are reported as median with IQR, and categorical variables as counts with percentages. Abbreviations: ALK, anaplastic lymphoma kinase; BM, brain metastasis; Dx, diagnosis; ECM, extracranial metastasis; EGFR, epidermal growth factor receptor; H&E, hematoxylin and eosin; IQR, interquartile range; KPS, Karnofsky Performance Status; NSCLC, non-small cell lung cancer; PD-L1, programmed death-ligand 1; SCLC, small cell lung cancer; TIL, tumor-infiltrating lymphocyte; TPS, tumor proportion score.

|  | NSCLC<br>Adenocarcinoma | NSCLC Non-Adenocarcinoma | SCLC | Overall |
| --- | --- | --- | --- | --- |
| Number of Patients | 74 | 18 | 11 | 103 |
| Age at BM Dx (years)<br>(median; IQR) | 66 (61 – 72) | 65.5 (58 – 71.5) | 69 (63 – 74) | 66 (60.5 – 72) |
| Sex |  |  |  |  |
| Male (%) | 27 (36%) | 9 (50%) | 7 (64%) | 43 (42%) |
| Female (%) | 47 (64%) | 9 (50%) | 4 (36%) | 60 (58%) |
| KPS at BM Dx (median;<br>IQR) | 75 (60 – 80) | 80 (70 – 80) | 70 (70 – 90) | 70 (60 – 85) |
| Num. BM at Dx (median;<br>IQR) | 2 (1 – 4) | 2 (1 – 4) | 5 (1 – 5.5) | 2 (1 – 5) |
| ECM Present at BM Dx (%) | 48 (65%) | 14 (78%) | 7 (64%) | 69 (67%) |
| EGFR+ (%) | 12 (16%) | 0 (0%) | 0 (0%) | 12 (12%) |
| ALK+ (%) | 1 (1%) | 0 (0%) | 0 (0%) | 1 (1%) |
| Number of Resected Lesions<br>(%) | 79 | 20 | 12 | 111 |
| Size of Dominant Lesion<br>(cm) (median; IQR) | 3.3 (2.5 – 4.0) | 3.5 (2.9 – 4.1) | 3.6 (1.6 – 4.5) | 3.3 (2.5 – 4.1) |
| PD-L1 Tumor Proportion<br>Score |  |  |  |  |
| 0% | 51 (65%) | 8 (40%) | 7 (58%) | 66 (59%) |
| 1-49% | 8 (10%) | 7 (35%) | 0 (0%) | 15 (14%) |
| ≥50% | 20 (25%) | 5 (25%) | 0 (0%) | 25 (23%) |

**Table 1. Baseline demographic, clinical, and histopathologic characteristics of the study cohort, stratified by histologic subtype.**
| Unavailable | 0 (0%) | 0 (0%) | 5 (42%) | 5 (5%) |
| --- | --- | --- | --- | --- |
| TIL Density |  |  |  |  |
| Absent (0) (%) | 31 (39%) | 7 (35%) | 0 (0%) | 38 (34%) |
| Low (1+) (%) | 35 (44%) | 10 (50%) | 11 (92%) | 56 (50%) |
| High (2+) (%) | 11 (14%) | 3 (15%) | 1 (8%) | 15 (14%) |
| Unavailable | 2 (3%) | 0 (0%) | 0 (0%) | 2 (2%) |

**Figure 1.**
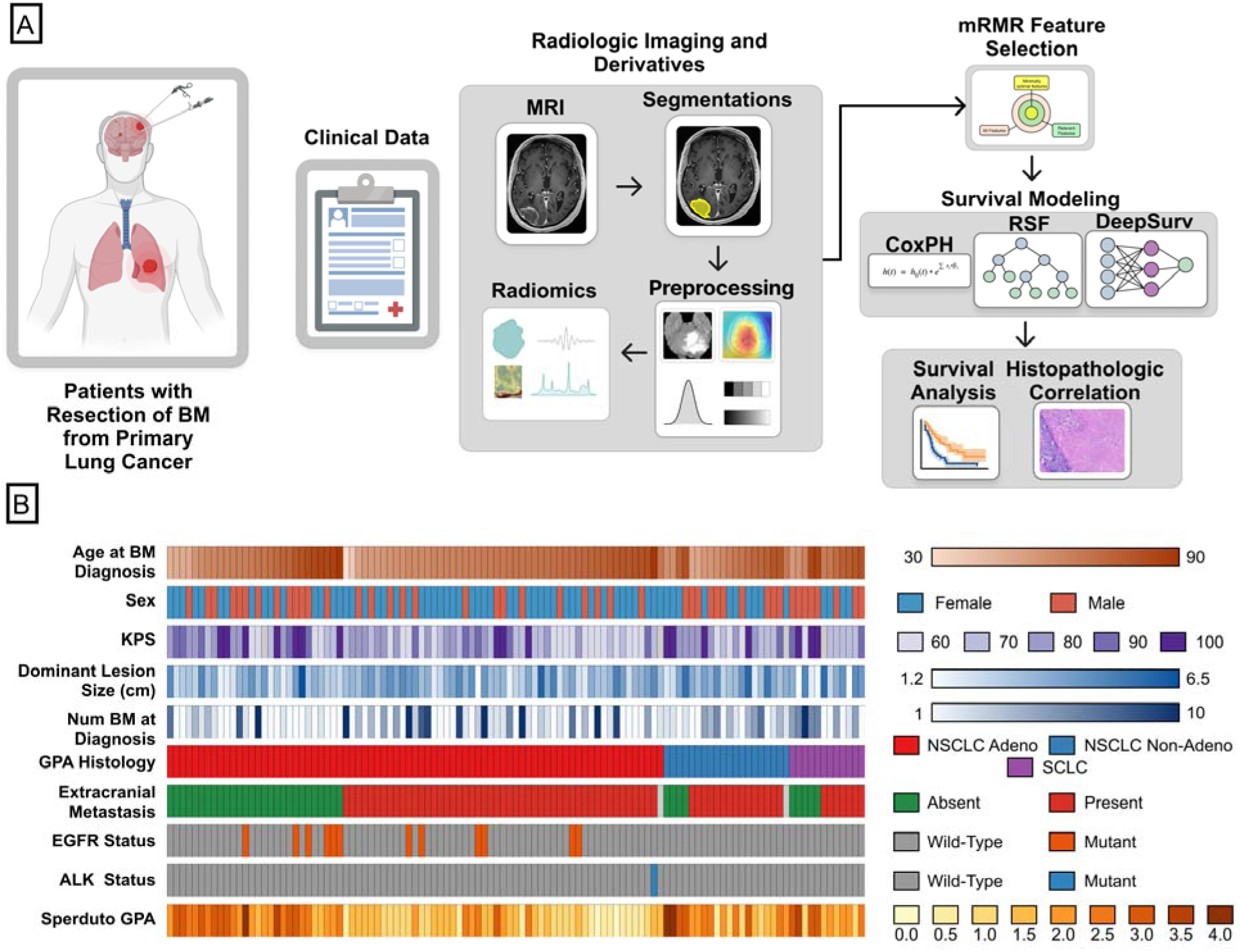
Study workflow and cohort characteristics. (A) Patients who underwent resection of BM from primary lung cancer were included in this retrospective study. Clinical variables were collected from the electronic health record. Pre-operative T1CE and FLAIR MRI sequences were retrieved from the imaging archive. The tumor core was manually segmented on T1CE images, and the whole tumor including peritumoral edema was segmented on FLAIR images. Images and segmentations underwent standardized preprocessing (brain extraction, N4 bias field correction, resampling, and intensity normalization), followed by radiomic feature extraction. Radiomic features were reduced using a variance threshold followed by mRMR selection. CoxPH, RSF, and DeepSurv models were compared. Patients were risk-stratified using the best-performing model, and survival was compared between risk groups using Kaplan–Meier analysis. Clinical and histopathologic characteristics were compared between high- and low-risk groups defined by the imaging-only model. (B) Heatmap of demographic, clinical, and molecular characteristics across the cohort, ordered by histology and extracranial metastasis status. Sperduto GPA scores range from 0 to 4, with higher scores indicating better prognosis. Abbreviations: ALK, anaplastic lymphoma kinase; BM, brain metastasis; CoxPH, Cox proportional hazards; EGFR, epidermal growth factor receptor; FLAIR, T2-weighted fluid-attenuated inversion recovery; GPA, Graded Prognostic Assessment; KPS, Karnofsky Performance Status; mRMR, minimal-redundancy-maximal-relevance; NSCLC, non-small cell lung cancer; RSF, random survival forest; SCLC, small cell lung cancer; T1CE, T1-weighted contrast-enhanced.

### Histopathologic Assessment

Fresh brain metastasis tissue was collected at the time of surgery and processed within one hour of collection. Specimens were formalin-fixed, paraffin-embedded, and stained with hematoxylin and eosin (H&E). H&E-stained sections underwent board-certified pathologist quality review to confirm adequate tumor cellularity and staining quality. Then, tumor-infiltrating lymphocyte (TIL) density was graded semi-quantitatively by board-certified pathologist evaluation on a three-tier scale: absent (0), low (1+), or high (2+), consistent with grading conventions used to classify TIL density in other solid tumors (20). For analysis, TIL grades were dichotomized as absent (0) or present (1+ or 2+). Programmed death-ligand 1 (PD-L1) expression was abstracted from the electronic health record (EHR) using the tumor proportion score (TPS), and analyzed at ≥1% and ≥50% thresholds.

### Outcomes of Interest

The primary outcome of interest was OS, defined as the interval from the date of surgical resection to death from any cause, censored at the time of last known follow-up. Secondary endpoints included CNS progression, defined as radiographic evidence of new or enlarging intracranial lesions on follow-up imaging, and development of LM metastasis based on cerebrospinal fluid cytology. Patients who did not experience a secondary outcome were censored at the date of death or last known follow-up. These outcomes were collected through EHR review.

### Image Preprocessing

Manual tumor segmentations were generated as described in our prior work (21). Briefly, the core tumor was defined as the region of contrast enhancement on T1CE imaging, while the whole tumor additionally included the surrounding peritumoral edema identified as FLAIR hyperintensity. Segmentation masks and images were exported as NIfTI files from the institutional picture archiving and communication system (PACS) for downstream processing.

A standardized preprocessing pipeline was then applied to all MRI volumes and segmentation masks prior to radiomic feature extraction. Brain extraction was performed using HD-BET to remove extracranial structures, including skull and scalp (22). Then, N4 bias field correction was applied to correct for spatial intensity inhomogeneities arising from magnetic field variation during image acquisition (23). To reduce variability in spatial resolution across scanners and acquisition protocols, all images were subsequently resampled to an isotropic voxel size of 1 × 1 × 1 mm³, using B-spline interpolation for image data and nearest-neighbor interpolation for segmentation masks (24). Finally, intensity normalization was performed using Nyúl histogram matching to reduce inter-scanner and inter-patient variability in MRI signal intensity distributions (25).

### Radiomic Feature Extraction

Radiomic features were extracted from three-dimensional tumor segmentations using PyRadiomics (version 3.1.0) in accordance with Image Biomarker Standardisation Initiative (IBSI) guidelines, employing B-spline interpolation and 25 intensity unit fixed-bin discretization (26). Features were computed from T1CE and FLAIR sequences using both original and wavelet-filtered images. This yielded 851 features per image: 14 shape features, plus 18 first-order statistics and 75 texture features computed on each of the original and eight wavelet-filtered images.

### Survival Models for Analysis

#### Survival Model Training

We compared three survival models—Cox Proportional Hazards (CoxPH), Random Survival Forest (RSF), and DeepSurv—to test how different assumptions about the feature-survival relationship affect performance. This range, from a linear baseline to a non-linear ensemble to a deep neural network, enabled us to evaluate how each modeling approach affected prognostic performance.

The CoxPH model serves as the linear statistical baseline (27). It estimates the hazard of an event as a function of covariates via a log-linear relationship, with coefficients estimated by maximizing the partial log-likelihood. CoxPH assumes proportional hazards across patients and is interpretable through its hazard ratios, making it well-suited for identifying individual prognostic features. However, it is limited by its inability to capture non-linear feature interactions.

The RSF model extends the CoxPH framework by accommodating non-linear relationships and higher-order interaction effects through an ensemble of survival decision trees (28). Each tree is grown on a bootstrap sample of the training data, and survival predictions are aggregated across the ensemble by consensus, yielding a more robust estimate of individual risk. RSF makes no parametric assumptions about the hazard function and can implicitly handle high-dimensional feature spaces. However, because each tree splits patients on one feature at a time, RSF represents continuous feature effects as discrete steps. In smaller cohorts, each terminal node also contains few patients, which can make individual risk estimates coarse and unstable.

DeepSurv employs a feedforward neural network trained using a Cox-based partial log-likelihood loss function, enabling the model to learn complex, high-dimensional non-linear feature interactions relevant to survival (17). The architecture consists of one to two fully connected hidden layers, with hyperparameters, including batch size, number of hidden layers, learning rate, and dropout rate, tuned via grid search. DeepSurv offers greater representational capacity than both CoxPH and RSF for capturing intricate feature relationships.

Consistent with methodological approaches used in prior radiomics-based prognostic and predictive modeling studies (29,30), all models were trained using five-fold cross-validation. Final model configurations were selected based on the highest validation concordance index (C-index).

#### Feature Selection

To reduce dimensionality prior to model training, radiomic features underwent a two-stage pipeline: low-variance features were first removed using a variance threshold of 1×10 to eliminate near-constant predictors, followed by the minimal-redundancy-maximal-relevance (mRMR) algorithm (31), which identifies features with maximal prognostic relevance while minimizing inter-feature redundancy. The top 40 radiomic features ranked by mRMR were retained for downstream modeling. This feature reduction process was conducted separately for each of the three study endpoints. Clinical covariates were not subject to this feature reduction.

#### Prognostic Value of Imaging Biomarkers

To determine whether MRI-derived radiomic features provided prognostic information beyond established clinical variables, survival models were trained using three distinct feature configurations: 1) clinical variables only; 2) imaging features only; and 3) combined clinical-imaging features. Clinical variables comprised the factors used in the lung cancer Graded Prognostic Assessment (GPA) (7): age, KPS, number of brain metastases, presence of extracranial metastases, EGFR/ALK status, PD-L1 expression, and histologic subtype. All three survival modeling frameworks (CoxPH, RSF, and DeepSurv) were evaluated under each feature configuration, yielding nine C-index values per outcome. This comparison was performed separately for each of the three study endpoints (OS, CNS progression, and LM metastasis) for a total of 27 C-index values across the study.

For each outcome, the best-performing model was selected from among all nine candidate models based on the highest C-index and used for prognostic risk stratification. Cases were ranked by predicted risk score and divided into thirds. The third with the highest predicted risk of the outcome was classified as high-risk, and the remaining two-thirds were classified as low-risk. Survival distributions between the resulting groups were compared using Kaplan–Meier analysis and the log-rank test. Risk groups used for correlative analyses are described in the corresponding sections below. We also evaluated whether model performance was consistent across clinically distinct patient subgroups. To assess this, C-index was calculated separately for subgroups based on histologic subtype and number of brain metastases at diagnosis.

#### Prognostic Value of Imaging Sequences

Prior radiomics studies of brain metastases and other CNS tumors differ in MRI sequences utilized. Some use only T1CE imaging to characterize the tumor core, while others add FLAIR to capture peritumoral signal changes (32,33). It is unclear whether FLAIR meaningfully improves prognostic performance. We therefore ran an ablation study, in which we removed the FLAIR-derived peritumoral edema features prior to feature reduction and compared models trained on T1CE features alone with models trained on the combined T1CE + FLAIR feature set for the OS endpoint.

### Interpretability and Imaging Feature Analysis

A key challenge with ML-based radiomic models is their reduced interpretability relative to traditional clinical models (34). Without insight into which feature patterns drive model predictions, it remains difficult to determine whether a model’s risk stratification reflects correlation with an underlying phenotype or a statistical artifact of the training data. We therefore performed two complementary analyses on the best-performing OS model, refit on the full cohort using the same hyperparameters. Feature contributions were quantified using SHapley Additive exPlanations (SHAP), reported as the mean absolute SHAP value for each input feature. To examine how the model organized patients, we extracted each patient’s penultimate-layer representation, computed pairwise Pearson correlations between patients, and performed unsupervised hierarchical clustering (average linkage, correlation distance). The two main clusters were compared for risk group, clinical input variables, and TIL and PD-L1 status using chi-square tests (Mann-Whitney U tests for continuous variables), and for OS using the log-rank test.

### Clinical Correlates of Imaging-Based Risk Groups

Established clinical prognostic models, such as the GPA, remain the standard for guiding treatment decisions, so benchmarking imaging-derived risk against them clarifies both its validity and potential added value of imaging features (7). To characterize the clinical correlates of imaging-derived risk, the distributions of GPA component variables were compared between high- and low-risk groups defined by the imaging-only DeepSurv model for each endpoint, using the same tertile-based definition described above. GPA components were dichotomized at their established GPA thresholds (7) and compared using chi-square tests. This analysis was intended to determine whether imaging-derived risk stratification recapitulates or provides information complementary to established clinical prognostic indices.

### Biologic Correlates of Imaging-Based Risk Groups

Because radiomic risk scores are derived through data-driven feature selection, it is not known whether they reflect underlying tumor or host biology. We therefore compared PD-L1 expression and TIL density between high- and low-risk groups defined by the imaging-only DeepSurv model for each endpoint. Associations were evaluated using chi-square tests.

### Statistical Analysis

Prognostic performance was assessed with the C-index, where 0.5 indicates no discrimination and 1.0 indicates perfect discrimination. To compare models and feature configurations, the C-index was computed on 1,000 bootstrap resamples of the patients within each cross-validation fold. Every model and configuration was evaluated on the same resamples, so they were compared with paired t-tests. Kaplan–Meier survival curves were constructed for imaging-derived high- and low-risk groups, with between-group differences assessed using the log-rank test. Associations between imaging-derived risk groups and categorical pathologic or clinical variables were evaluated using chi-square tests. A two-sided *p*-value < 0.05 was considered statistically significant for all analyses.

### Code Availability

All analyses were conducted in Python. Code for preprocessing, feature extraction, model development, and evaluation is publicly available on the Aneja Lab’s GitHub profile (https://github.com/Aneja-Lab-Yale/Aneja-Lab-Public-Lung-Brain-Mets-MRI-Biomarker).

### Data Availability

Pre-operative MRI sequences (T1CE and FLAIR), radiomic feature files, segmentation masks, clinical metadata, and digitized pathology images are publicly accessible through The Cancer Imaging Archive (TCIA) (https://doi.org/10.7937/k0sm-y874). Additional data may be made available upon reasonable request to the corresponding author, subject to Yale University Institutional Review Board and data use agreement requirements.

## Results

### Study Cohort

The cohort included 111 resected brain metastases from 103 patients with lung cancer (Table 1). Most lesions were adenocarcinoma (71%), followed by non-adenocarcinoma non-small cell lung cancer (NSCLC; 18%) and small cell lung cancer (SCLC; 11%). Patients had a median age of 66 years (interquartile range [IQR], 60.5–72) at brain metastasis diagnosis, and 58% were female. The median KPS was 70 (IQR, 60–85), the median number of brain metastases was 2 (IQR, 1–5), and 67% of patients had extracranial metastases at brain metastasis diagnosis. The median dominant lesion diameter was 3.3 cm (IQR, 2.5–4.1). EGFR mutations and ALK alterations occurred only in adenocarcinoma, in 16% and 1% of these patients, respectively. PD-L1 expression (TPS ≥1%) was present in 35% of adenocarcinoma and 60% of non-adenocarcinoma NSCLC lesions. All SCLC lesions had TILs present, mostly at low density (92%), compared to 58–65% of NSCLC lesions.

### Prognostic Value of Imaging Biomarkers

For OS, imaging-derived radiomic features improved prognostic performance beyond clinical variables alone across all survival model types (Figure 2A). Clinical-only models yielded C-index values of 0.643, 0.621, and 0.680 for CoxPH, RSF, and DeepSurv, respectively. Imaging-only models yielded C-index values of 0.684, 0.652, and 0.719 for CoxPH, RSF, and DeepSurv, which were significantly higher than the clinical-only models (all *p <* 0.001). Combined clinical-imaging models achieved the highest C-index values—0.704 for CoxPH, 0.658 for RSF, and the clinical-imaging DeepSurv model demonstrated the highest C-index value overall of 0.728. The clinical-imaging models exhibited significantly higher C-index values than the clinical-only models across all survival modeling frameworks (all *p* < 0.001). The clinical-imaging models for CoxPH and DeepSurv also had significantly higher C-index values than the imaging-only models (both *p <* 0.001), while for RSF, the clinical-imaging and imaging-only models were comparable (*p* = 0.076). When patients were stratified into low- and high-risk groups using prognostic scores derived from the clinical-imaging DeepSurv model, Kaplan–Meier analysis demonstrated a significant difference in OS between groups, with a median OS of 26.7 months in the low-risk cohort versus 9.1 months in the high-risk cohort (*p* < 0.001; Figure 2B).

**Figure 2.**
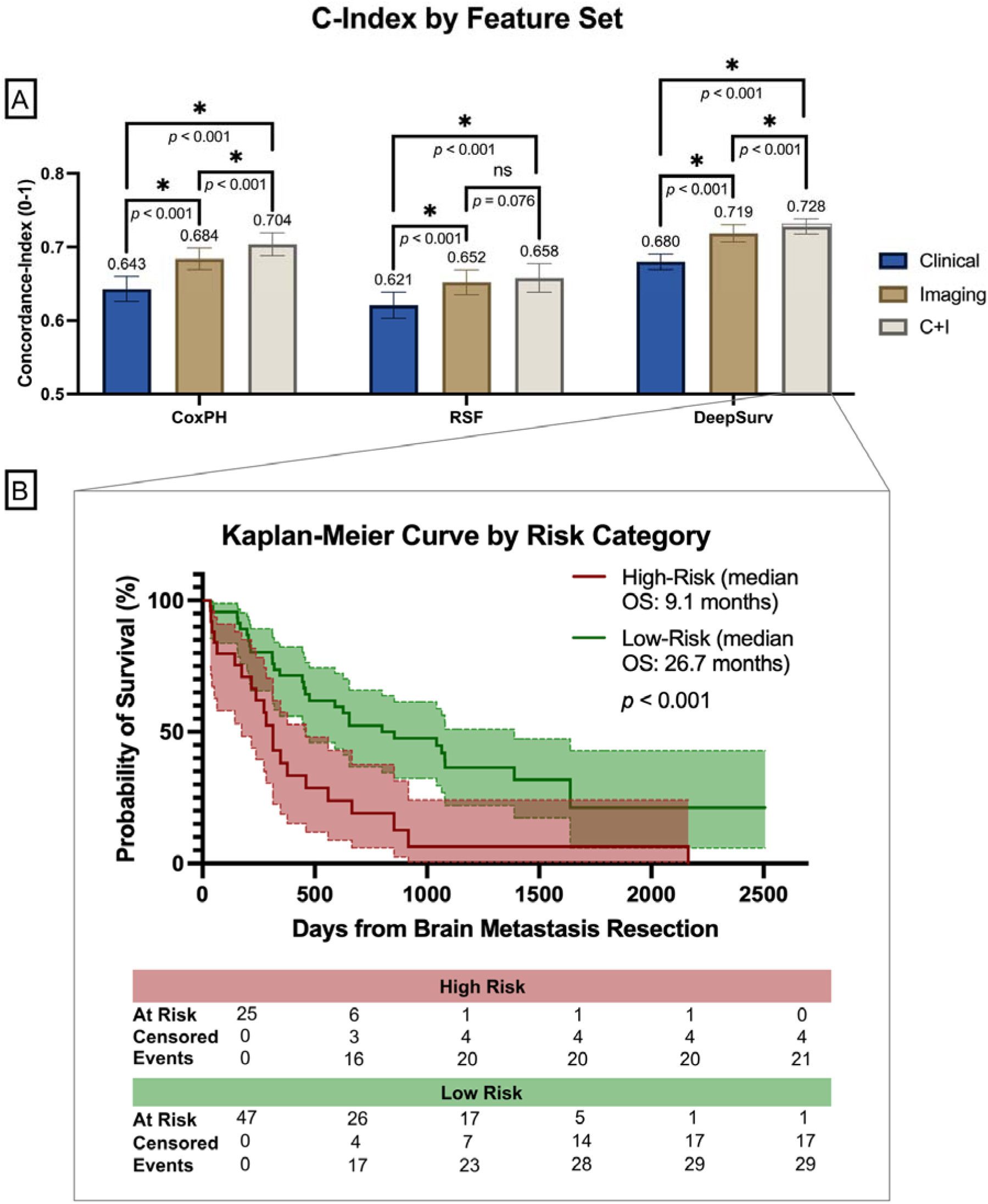
Overall survival model performance by feature set and Kaplan–Meier analysis of risk groups. (A) Concordance index (C-index) of CoxPH, RSF, and DeepSurv models using clinical variables alone, imaging features alone, or combined clinical and imaging (C+I) features. Error bars represent 95% bootstrap confidence intervals; brackets show pairwise comparisons. \**p* < 0.05; ns, not significant. (B) Kaplan–Meier curves for risk groups defined by the best-performing model, the clinical-imaging DeepSurv model. Patients in the highest third of predicted risk were classified as high-risk and those in the remaining two-thirds were labeled as low-risk. Numbers at risk, censored, and events are shown below the plot. The *p*-value was calculated using the log-rank test. Abbreviations: C+I, combined clinical and imaging; C-index, concordance index; CoxPH, Cox proportional hazards; RSF, random survival forest.

Prognostic performance of the combined clinical-imaging DeepSurv OS model varied across patient subgroups; however, this variation was not statistically significant. C-index values were 0.720 for adenocarcinoma NSCLC, 0.624 for non-adenocarcinoma NSCLC, and 0.857 for SCLC (Figure S1A). Pairwise comparisons did not show any significant differences between performance on adenocarcinoma and non-adenocarcinoma NSCLC (*p =* 0.331), adenocarcinoma and SCLC (*p* = 0.263), or non-adenocarcinoma and SCLC (*p* = 0.122). Prognostic performance was similarly comparable (*p =* 0.886) between patients with two or fewer brain metastases at diagnosis (C-index 0.713) and those with more than two (C-index 0.700; Figure S1B).

For the CNS progression outcome, imaging-derived features improved prognostic performance beyond clinical variables alone across all three model types (Figure S2A), though risk-group stratification did not reach statistical significance (Figure S2B). Clinical-only models yielded C-index values of 0.435, 0.352, and 0.560 for CoxPH, RSF, and DeepSurv, respectively. Imaging-only models improved performance to 0.564, 0.576, and 0.701, and combined clinical-imaging models achieved C-index values of 0.520, 0.568, and 0.695, respectively. When patients were stratified into low- and high-risk groups, Kaplan–Meier analysis showed a median time to CNS progression of 7 months in the high-risk group versus 15 months in the low-risk group; however, this difference did not reach statistical significance (*p* = 0.069).

For the LM metastasis outcome, the incorporation of imaging features improved prognostic performance for the CoxPH and DeepSurv models (Figure S3A). Clinical-only models yielded C-index values of 0.594, 0.633, and 0.685 for CoxPH, RSF, and DeepSurv, respectively. Imaging-only models demonstrated C-index values of 0.700, 0.584, and 0.751, and combined clinical-imaging models achieved C-index values of 0.660, 0.595, and 0.756, respectively. LM progression risk-group stratification using the clinical-imaging DeepSurv model reached statistical significance, with a median time to progression not reached in the low-risk group compared to 22.0 months in the high-risk group (*p* = 0.003; Figure S3B).

### Prognostic Value of Imaging Sequences

Adding FLAIR-derived peritumoral edema features to T1CE improved OS prediction in the CoxPH and DeepSurv models, though not in RSF (Figure 3). In a modality ablation study comparing our primary T1CE+FLAIR models against T1CE-only models, adding FLAIR raised the C-index of imaging-only models from 0.663 to 0.684 for CoxPH (*p* < 0.001) and from 0.705 to 0.719 for DeepSurv (*p* < 0.001), while RSF showed no significant change (0.665 vs. 0.652, *p* = 0.520). The same pattern held in combined clinical-imaging models, where FLAIR raised the C-index from 0.668 to 0.704 for CoxPH (*p* < 0.001) and from 0.702 to 0.728 for DeepSurv (*p* < 0.001), with no significant difference for RSF (0.649 vs. 0.658, *p* = 0.062).

**Figure 3.**
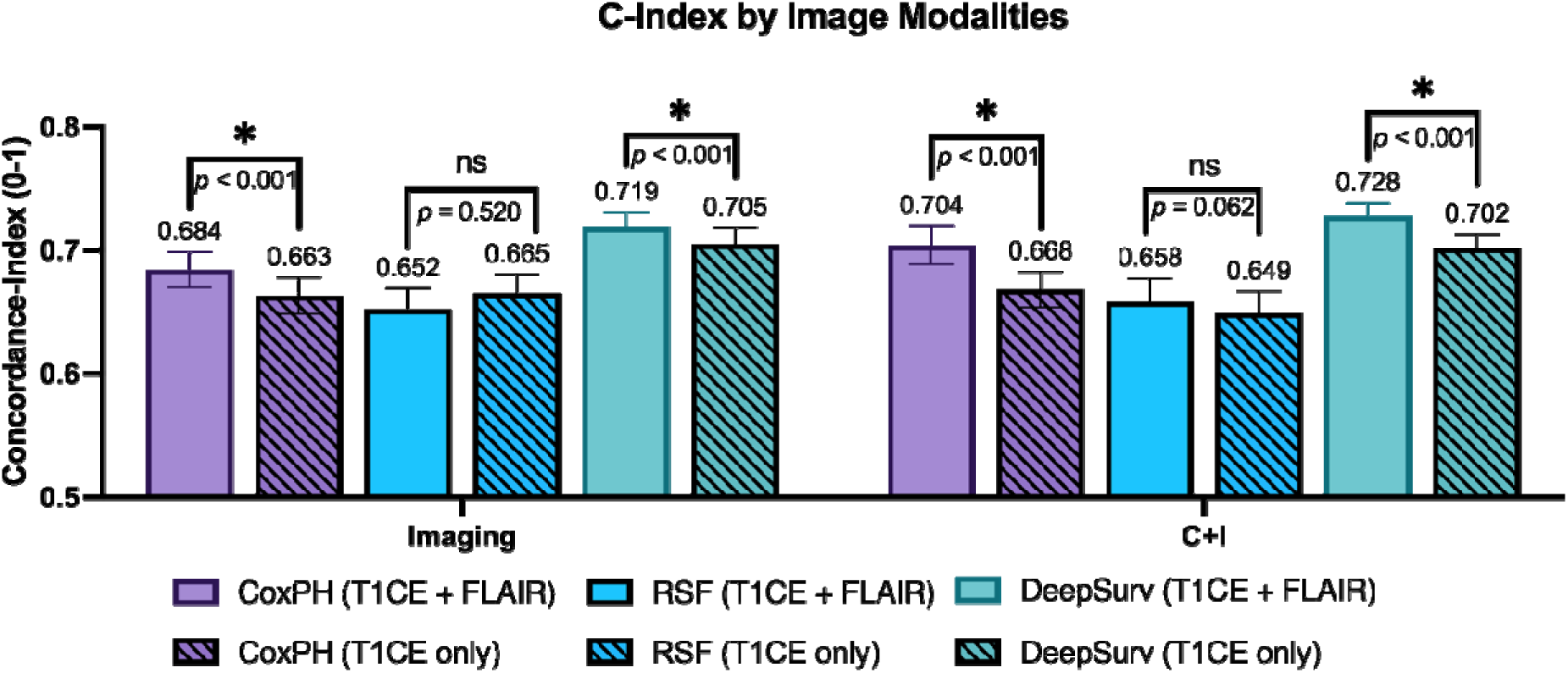
Overall survival model performance by MRI sequence. C-index of models trained on T1CE features alone versus combined T1CE and FLAIR features, shown for imaging-only and combined clinical-imaging (C+I) models. Adding FLAIR significantly improved performance for CoxPH and DeepSurv but not RSF. Error bars represent 95% bootstrap confidence intervals; brackets show pairwise comparisons. \**p* < 0.05; ns, not significant. Abbreviations: C+I, combined clinical and imaging; C-index, concordance index; CoxPH, Cox proportional hazards; FLAIR, T2-weighted fluid-attenuated inversion recovery; RSF, random survival forest; T1CE, T1-weighted contrast-enhanced.

### Interpretability and Imaging Feature Analysis

Radiomic features dominated model attributions, accounting for 15 of the 20 most influential features (Figure 4B); the top three were maximum 2D diameter, age, and zone entropy. Unsupervised clustering of the model’s learned representations identified two patient clusters that differed markedly in predicted risk (7% vs. 55% high-risk, *p* < 0.001; Figure 4A). Compared with the high-risk cluster, the low-risk cluster had longer OS (median 53.8 vs. 9.3 months, log-rank *p* < 0.001), more TIL-present tumors (79% vs. 55%, *p* = 0.036), more PD-L1 ≥ 50% expression (43% vs. 11%, *p* = 0.002), and fewer patients with extracranial metastasis (53% vs. 83%, *p* = 0.008). Histology, PD-L1 ≥1%, number of lesions, and age did not differ significantly (all *p* > 0.05).

**Figure 4.**
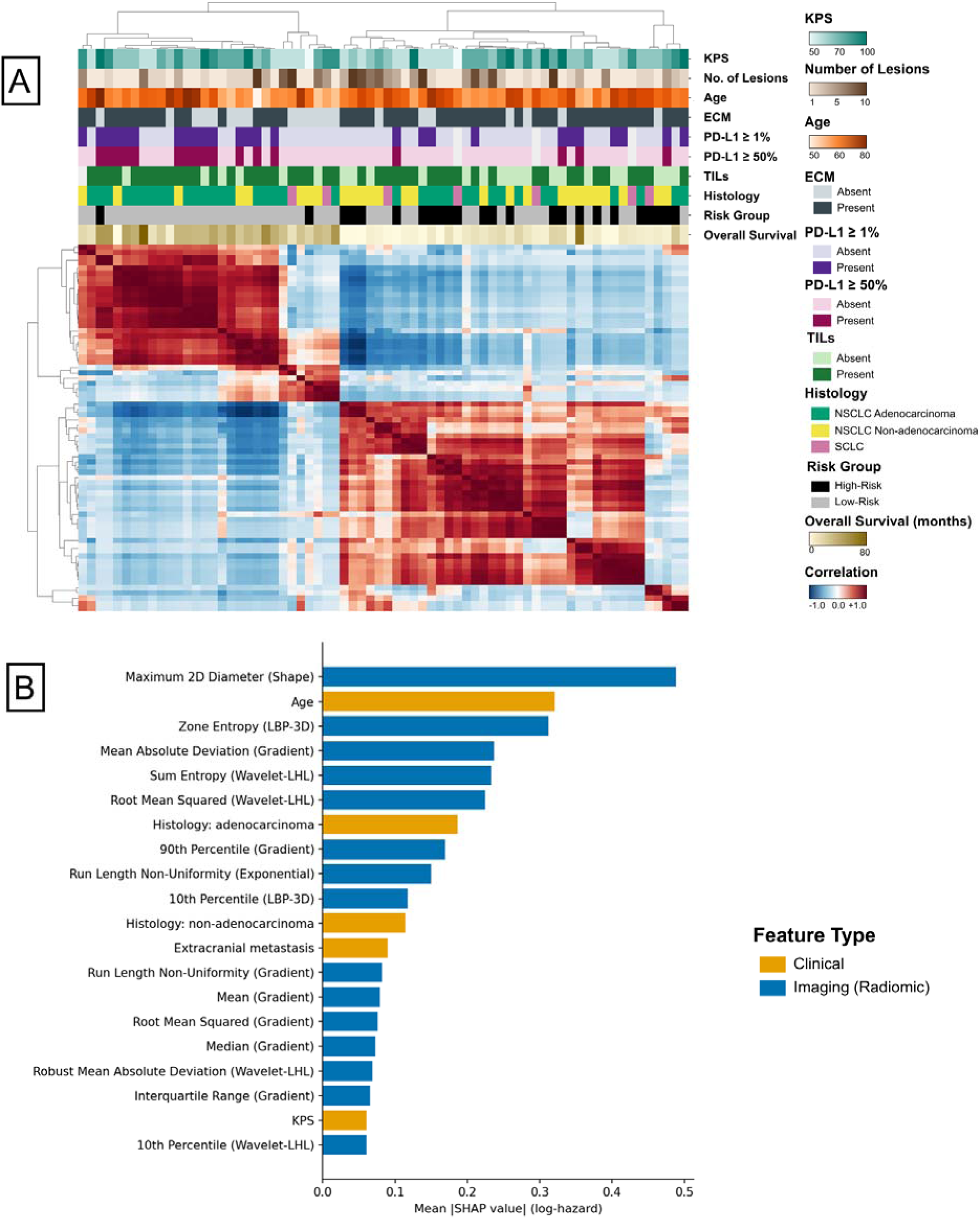
Interpretability of the clinical-imaging DeepSurv overall survival model. (A) Unsupervised hierarchical clustering of patient-by-patient Pearson correlations between penultimate-layer model representations (average linkage, correlation distance) identified two patient clusters. Annotation tracks show model-derived risk group, overall survival (for reference; not a model input), and clinical input variables. (B) The 20 most influential model features ranked by mean absolute SHAP value, colored by feature type. Both analyses used a model fit on the full cohort. Abbreviations: ECM, extracranial metastasis; KPS, Karnofsky Performance Status; PD-L1, programmed death-ligand 1; SHAP, SHapley Additive exPlanations; TIL, tumor-infiltrating lymphocyte.

### Clinical Correlates of Imaging-Based Risk Groups

To assess whether imaging-defined risk groups reflected established clinical prognostic factors, we compared GPA components between high-risk and low-risk patients defined by the imaging-only DeepSurv OS model (Figure 5A). No individual GPA component differed significantly between groups, including KPS, number of brain metastases at diagnosis, age at diagnosis, and extracranial metastatic disease (all *p* > 0.05). Composite GPA showed a non-significant trend toward lower scores in the high-risk group (96% vs. 79% with GPA 0–2.5, *p* = 0.064). Similarly, for the CNS progression and LM metastasis endpoints, no clinical variable differed significantly between imaging-only risk groups (all *p* > 0.05).

**Figure 5.**
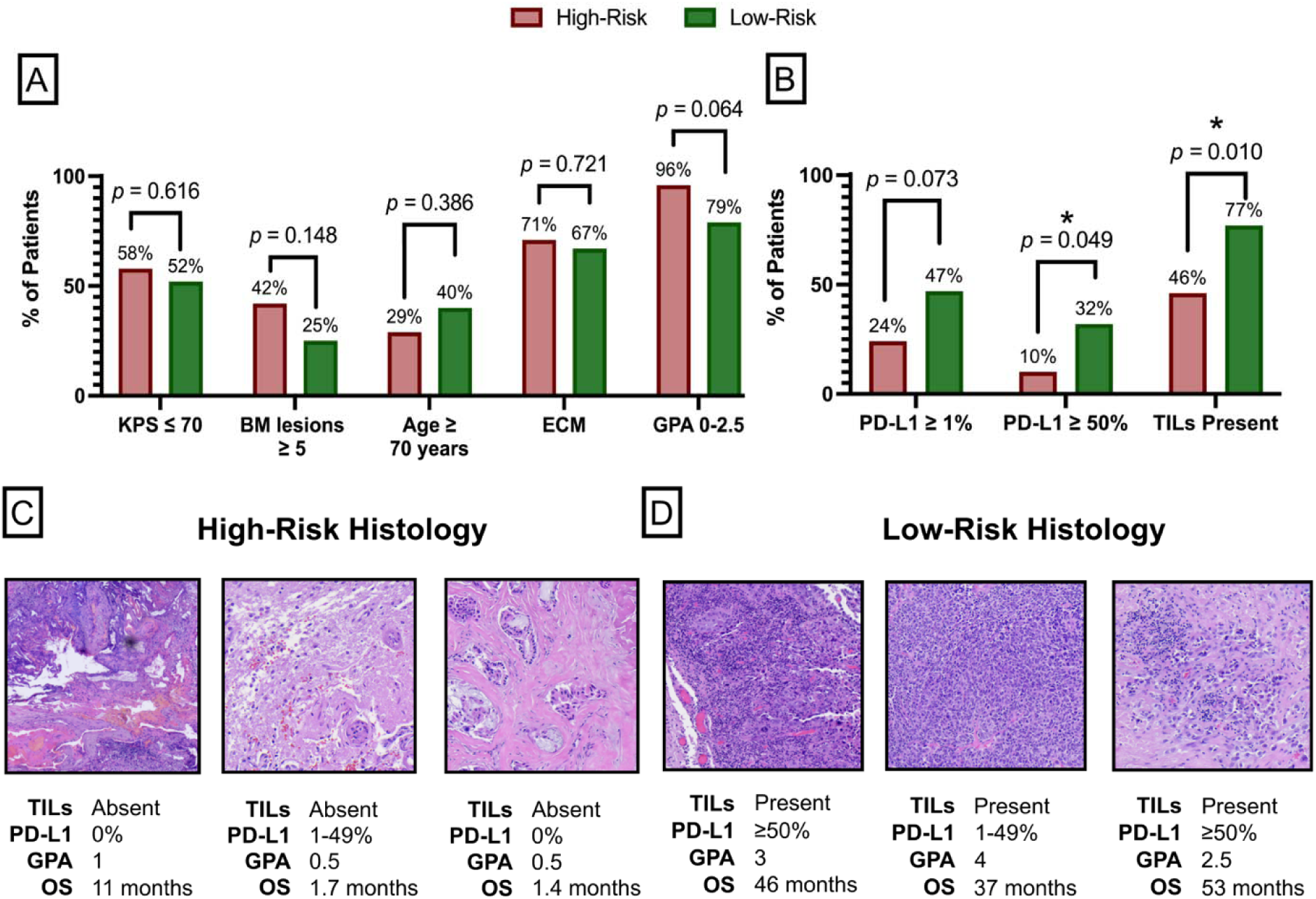
Clinical and histopathologic correlates of imaging-defined risk groups. Patients were divided into high-risk (highest third of predicted risk) and low-risk (remaining two-thirds) groups by the imaging-only DeepSurv OS model, which uses no clinical or pathologic inputs. (A) Proportion of patients with the unfavorable value of each GPA component (KPS ≤70, ≥5 BM at diagnosis, age ≥70 years, ECM) and with a composite GPA score of 0–2.5 (range 0–4; lower scores indicate worse prognosis). No component differed significantly between groups. (B) Proportion of patients with PD-L1 TPS ≥1% and ≥50%, obtained from clinical pathology reports, and with TILs present on matched H&E-stained resection specimens, graded by a board-certified pathologist. High-risk patients were significantly less likely to have PD-L1 ≥50% and TIL-present tumors. (C, D) Representative H&E-stained sections from three high-risk (C) and three low-risk (D) patients, with TIL status, PD-L1 TPS, GPA score, and OS shown for each. *P*-values were calculated using chi-square tests; \**p* < 0.05. Abbreviations: BM, brain metastasis; ECM, extracranial metastasis; GPA, Graded Prognostic Assessment; H&E, hematoxylin and eosin; KPS, Karnofsky Performance Status; OS, overall survival; PD-L1, programmed death-ligand 1; TIL, tumor-infiltrating lymphocyte; TPS, tumor proportion score.

### Biologic Correlates of Imaging-Based Risk Groups

For OS, high-risk patients defined by the imaging-only DeepSurv model were less likely than low-risk patients to have TIL-present tumors (46% vs. 77%, *p* = 0.010) and PD-L1 ≥50% expression (10% vs. 32%, *p* = 0.049), with a similar trend at the ≥1% threshold (24% vs. 47%, *p* = 0.073; Figure 5B). Representative histopathologic specimens for high-risk and low-risk patients are shown in Figures 5C and 5D, respectively. For CNS progression, imaging-only high-risk patients were also less likely to have PD-L1 ≥ 50% (11% vs. 37%, *p* = 0.017), with a trend at the ≥ 1% threshold (26% vs. 48%, *p* = 0.057), while TIL-present status did not differ significantly (56% vs. 62%, *p* = 0.607). For LM metastasis, PD-L1 status and TIL density did not differ significantly between imaging-only risk groups (all *p* > 0.05).

## Discussion

Brain metastases from lung cancer are a major clinical challenge, and existing clinical prognostic models only partly capture the variation in patient outcomes. In this study, we developed and validated ML survival models that combine clinical variables with MRI radiomic features to improve risk stratification for patients with brain metastases.

Adding MRI radiomic features to clinical prediction models improved survival prediction across three different survival model types. Radiomics converts standard-of-care imaging into quantitative features that capture texture and intensity patterns across the whole lesion, including tumor heterogeneity that may not be visually apparent. This is especially useful in brain metastases, where features are extracted from imaging already obtained during diagnostic workup and therefore add no delay to time-sensitive treatment decisions. Combining clinical and radiomic features has improved outcome prediction in other cancers. In NSCLC, combining CT body radiomics with clinical staging improved survival prediction over either alone (35), with similar gains also in head-and-neck (36) and urothelial carcinomas (37). In NSCLC brain metastases specifically, Liao et al. found combined clinical-imaging models outperformed clinical models alone for local control and survival after radiosurgery (38), and Zhang et al. found that a nomogram combining CT radiomic scores with KPS and sex predicted OS after whole-brain radiotherapy (39).

Our work builds on prior studies showing prognostic value of MRI for brain metastases by evaluating three clinically relevant endpoints across three different survival model types. Pre-treatment T1CE MRI features have been shown to independently predict local tumor control following Gamma Knife radiosurgery (33), and MRI features have predicted brain metastasis velocity and survival in other radiosurgery cohorts (40). These studies focused mainly on intracranial disease control after radiosurgery. We instead modeled OS after resection as the primary endpoint, with CNS and LM progression as secondary endpoints. Unlike OS, which is strongly influenced by extracranial disease, CNS and LM progression may more directly reflect the intracranial disease captured on brain MRI. Identifying patients at risk for LM disease after resection is especially important, given its poor prognosis and limited treatment options (41). In our cohort, imaging features improved prediction of both secondary endpoints, and imaging-derived risk groups differed significantly in time to LM progression. Prior radiomic prognostic studies have also typically relied on a single modeling approach, making it difficult to separate the contribution of imaging features from the choice of model. We therefore evaluated each endpoint across three different modeling frameworks. Imaging features improved prediction in nearly all endpoint–model combinations, including all three OS models, suggesting that their prognostic value is largely consistent across modeling approaches.

We also extend prior radiomic studies of brain metastases by demonstrating the prognostic value of peritumoral FLAIR imaging alongside T1CE. Most prior studies have characterized only the enhancing tumor on T1CE, although the peritumoral region contains vasogenic edema and, in some lesions, infiltrating tumor cells that may reflect more invasive behavior (42). In our modality ablation study, adding FLAIR features to T1CE improved OS prediction in the CoxPH and DeepSurv models for both imaging-only and combined clinical-imaging configurations, while RSF showed no significant change. Because FLAIR is acquired routinely in brain metastasis imaging protocols, these features can be incorporated without additional imaging.

Risk groups from the best-performing model differed significantly in OS. To understand what the imaging features capture independently, we examined risk groups from the imaging-only model, which uses no clinical or pathologic inputs. These groups did not differ significantly in any GPA component, although most components trended toward less favorable values in the high-risk group. This suggests that the radiomic signature is not simply a proxy for established clinical risk factors.

Our study is among the first to better understand the relationship between MRI radiomic features and brain metastasis biology. Because every lesion in our cohort was resected, we could compare imaging-defined risk directly with histopathology from the same lesion. High-risk patients defined by imaging alone were less likely to have TIL-present tumors and PD-L1 ≥50% expression, suggesting that the radiomic signature reflects, in part, the tumor immune microenvironment. Both markers point to the same underlying phenotype. PD-L1 expression on tumor cells is largely induced by interferon-γ from infiltrating cytotoxic T cells (43,44), so PD-L1 positivity often marks a lesion with an active immune response, while PD-L1 negativity more often reflects an immunologically cold tumor. Consistent with this, PD-L1 expression correlates strongly with TIL density across large brain metastasis cohorts (45) and has been associated with survival and intracranial disease control in NSCLC brain metastases (46). The same immune biology may explain why FLAIR features added prognostic value. Peritumoral FLAIR hyperintensity reflects vasogenic edema arising from vascular endothelial growth factor (VEGF)-mediated disruption of endothelial tight junctions (47), and TILs are a source of VEGF and inflammatory cytokines in the metastatic niche (48,49). Berghoff et al. observed this relationship directly, with denser TIL infiltration correlating with both more extensive peritumoral edema and longer OS in patients with brain metastases (49). Together, these findings suggest that the developed imaging biomarker may offer a noninvasive window into tumor immune phenotype.

Limited interpretability remains a barrier to clinical adoption of ML-based radiomic models (34). We therefore examined which features contributed most to predictions in the best-performing clinical-imaging OS model. Radiomic features accounted for 15 of the 20 most influential features, suggesting that model predictions drew heavily on imaging features. Maximum 2D diameter, a measure of lesion size, was the most influential feature, consistent with prior studies identifying tumor size and volume as independent predictors of survival in brain metastases (50). Tumor size is not currently included in the GPA (7), highlighting one way radiomic features may complement established clinical indices. Age, a current GPA component, ranked second, followed by zone entropy, a texture feature that captures intratumoral heterogeneity, which has been associated with prognosis across cancers (11). Together with the histopathologic correlation above, this analysis provides further evidence that our model uses biologically meaningful imaging features not captured by current clinical indices.

Our study has limitations. The modest sample size may limit generalizability. This reflects our focus on a surgical cohort with matched histopathology, which allowed imaging features to be compared directly with tissue from the same lesion. To reduce the risk of overfitting, we used survival models designed for censored data, five-fold cross-validation with bootstrapping, and dimensionality reduction of radiomic features before training. Because patients selected for resection typically have limited intracranial disease, our findings may not extend to patients with many metastases, although sensitivity analysis showed consistent performance in patients with fewer and more numerous brain metastases. Our models were also trained only on patients with lung cancer and may not apply to brain metastases from other primary tumors. As a retrospective study, our analysis may be subject to selection bias, and differences in MRI acquisition parameters may add variance to radiomic measurements, which we addressed through standardized preprocessing, including resampling, bias field correction, and intensity normalization. No standardized TIL scoring system exists for brain metastases, so TIL density was graded semi-quantitatively by board-certified pathology review using a three-tier scale analogous to those used in melanoma (20), and grades were dichotomized as absent or present for analysis. Validation in larger, independent cohorts as well as prospective evaluation are ongoing.

In conclusion, ML models using MRI features improved survival prediction beyond clinical variables in patients with surgically resected brain metastases from lung cancer. Imaging-defined risk groups differed in TIL presence and PD-L1 expression on matched histopathology, suggesting that these features may reflect the local inflammatory response and tumor immune microenvironment. Current prognostic models for brain metastases rely on clinical variables alone. Combined clinical-imaging biomarkers could address this gap in precision prognostication and help inform decisions about surgical resection, radiotherapy, and systemic therapy at the time of diagnosis.

## Acknowledgements

Research reported in this publication was supported by the Richard K. Gershon Endowed Medical Student Research Fellowship.

