## Supplementary Figures for "MRI Biomarkers for Brain Metastases in Lung Cancer Using Machine Learning"

**Supplemental Figures**

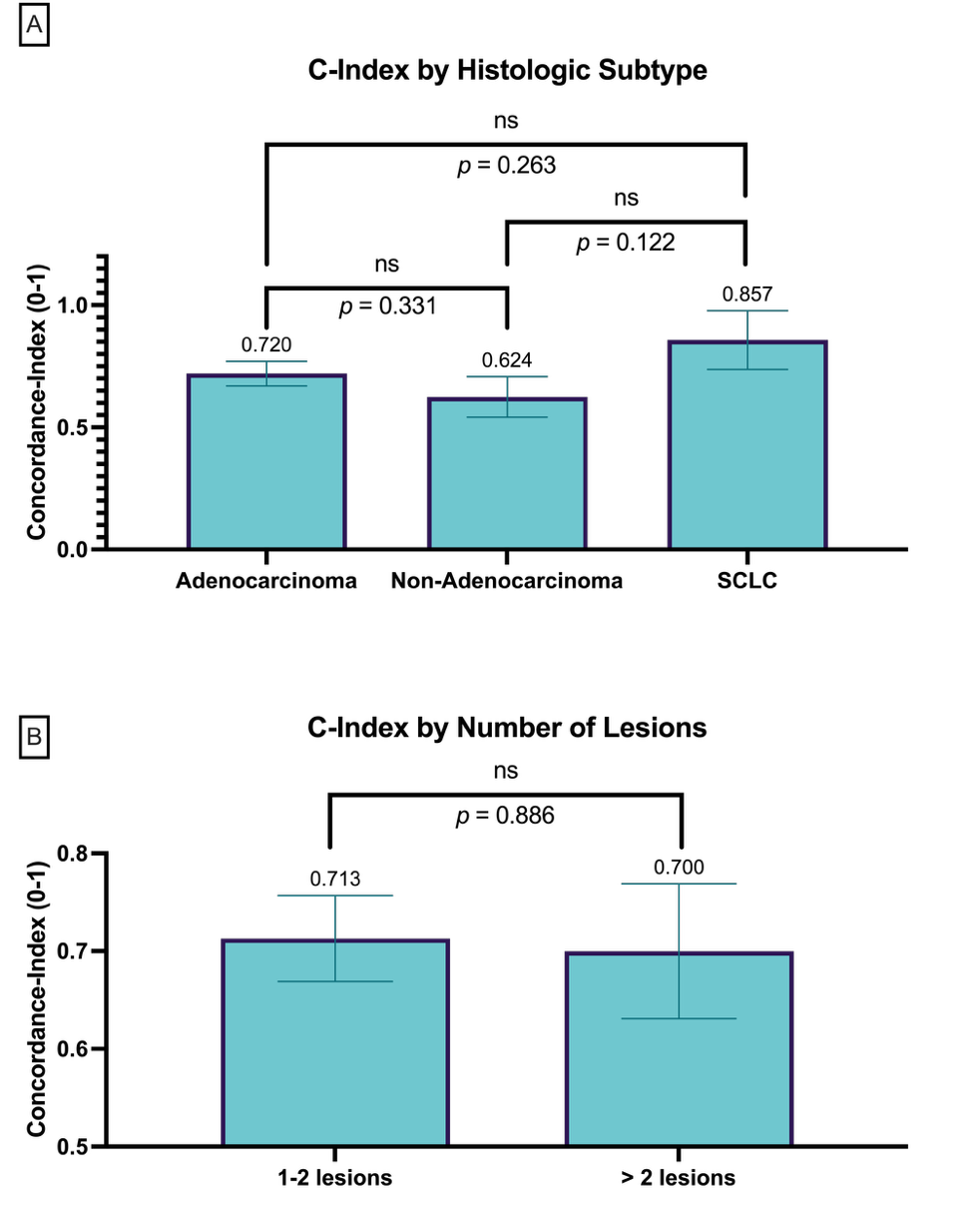

**Supplementary Figure S1. Overall survival model performance across clinical subgroups.** C-index of the combined clinical-imaging DeepSurv model, stratified by (A) histologic subtype (adenocarcinoma NSCLC, non-adenocarcinoma NSCLC, and SCLC) and (B) number of brain metastases at diagnosis (1-2 vs. >2). Performance did not differ significantly between subgroups. Error bars represent 95% bootstrap confidence intervals; *p*-values are from pairwise comparisons. ns, not significant. Abbreviations: C-index, concordance index; NSCLC, non-small cell lung cancer; SCLC, small cell lung cancer.

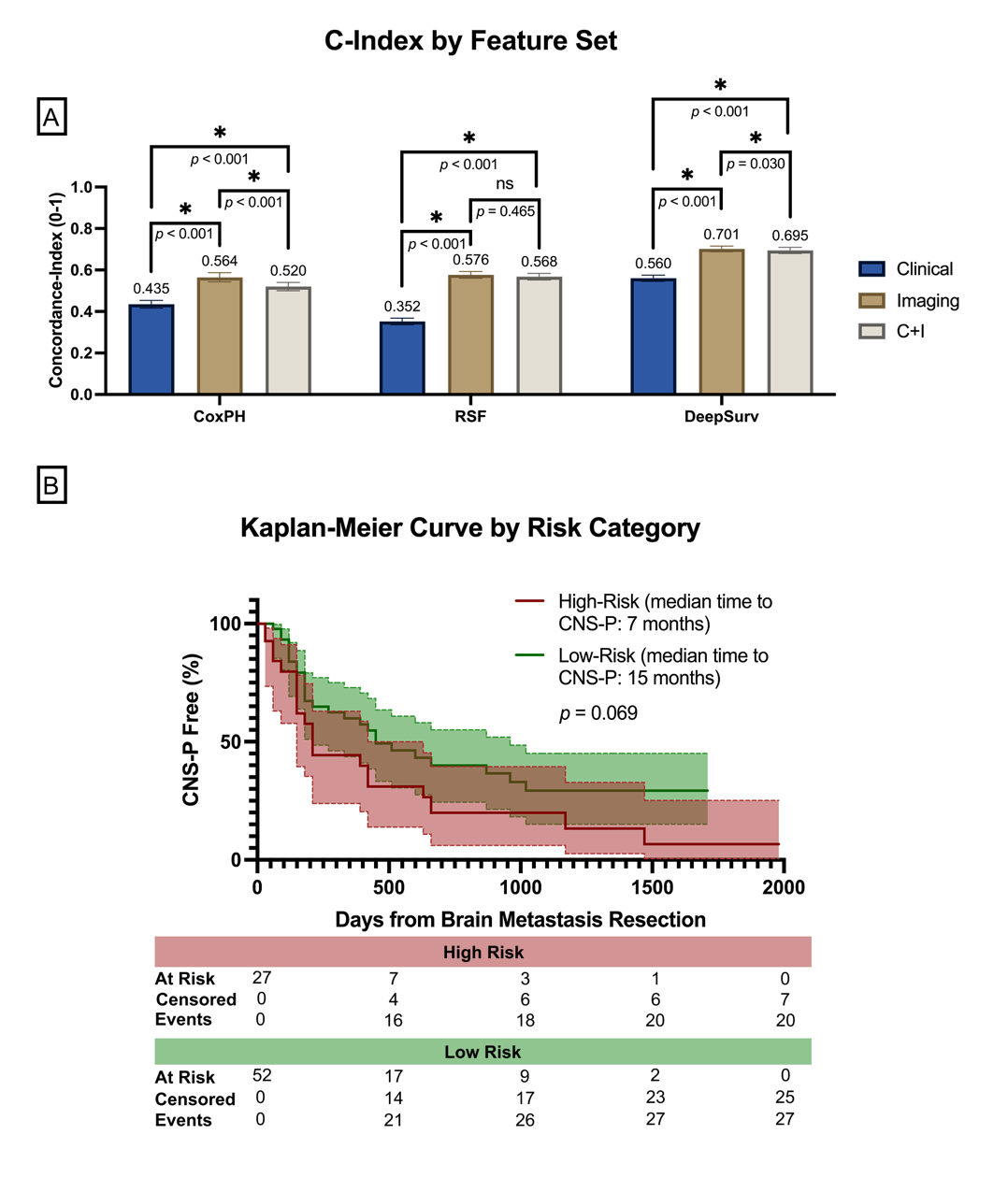

**Supplementary Figure S2.** **CNS progression model performance by feature set and Kaplan–Meier analysis of risk groups.** (A) C-index of CoxPH, RSF, and DeepSurv models for CNS progression using clinical variables alone, imaging features alone, or combined clinical and imaging (C+I) features. Error bars represent 95% bootstrap confidence intervals; brackets show pairwise comparisons. **p* < 0.05; ns, not significant. (B) Kaplan–Meier curves from best-performing CNS progression survival model. Patients in the highest third of predicted risk were classified as high-risk and the remaining two-thirds as low-risk. Numbers at risk, censored, and events are shown below the plot. The *p*-value was calculated using the log-rank test. Abbreviations: C+I, combined clinical and imaging; C-index, concordance index; CNS, central nervous system; CoxPH, Cox proportional hazards; RSF, random survival forest.

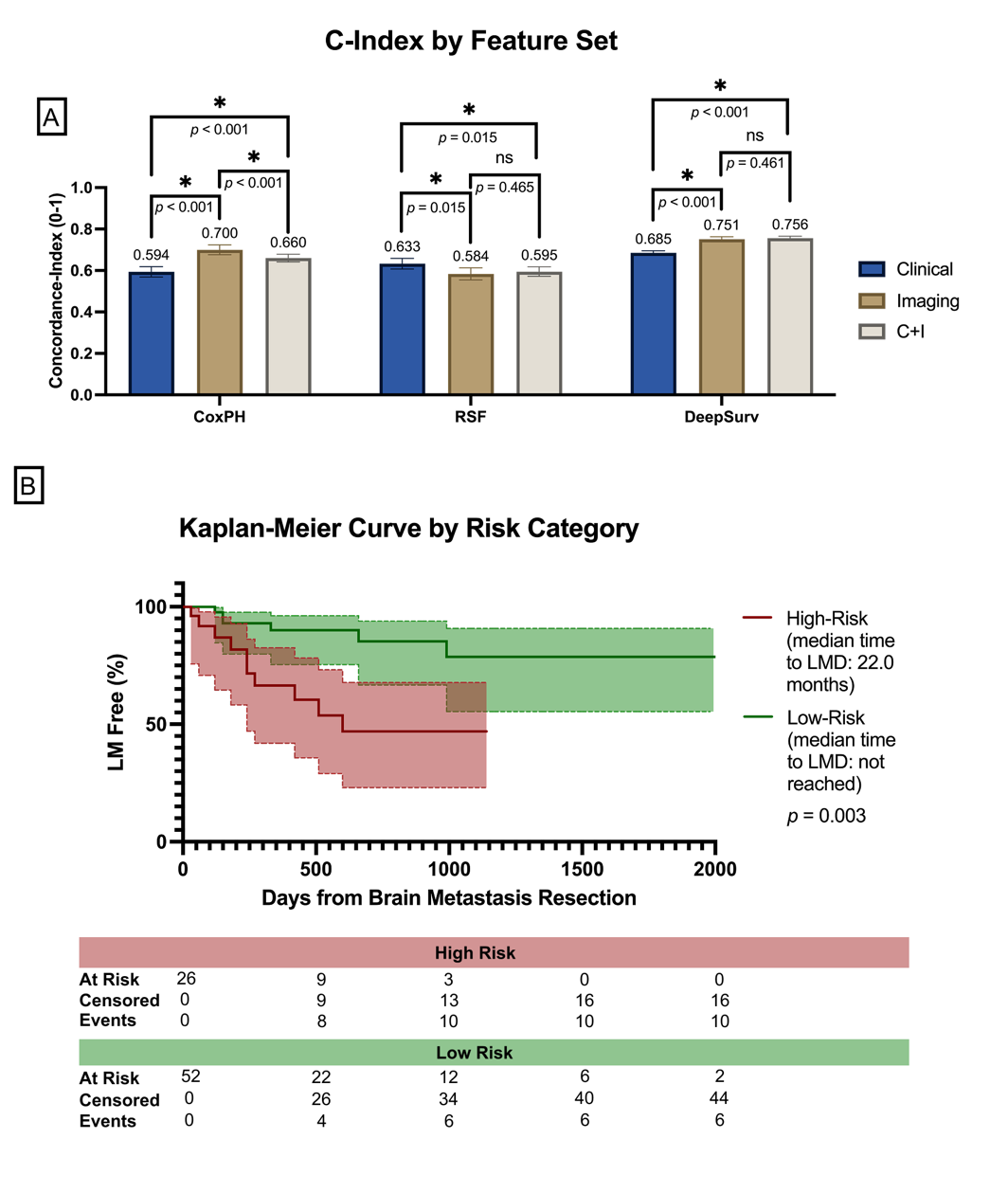

**Supplementary Figure S3.** **Leptomeningeal metastasis model performance by feature set and Kaplan–Meier analysis of risk groups.** (A) C-index of CoxPH, RSF, and DeepSurv models for leptomeningeal metastasis (LM) using clinical variables alone, imaging features alone, or combined clinical and imaging (C+I) features. Error bars represent 95% bootstrap confidence intervals; brackets show pairwise comparisons. **p* < 0.05; ns, not significant. (B) Kaplan–Meier curves from the best-performing LM survival model. Patients in the highest third of predicted risk were classified as high-risk and the remaining two-thirds were classified as low-risk. Numbers at risk, censored, and events are shown below the plot. The *p*-value was calculated using the log-rank test. Abbreviations: C+I, combined clinical and imaging; C-index, concordance index; CoxPH, Cox proportional hazards; LM, leptomeningeal metastasis; RSF, random survival forest.
